# Internet-based Cognitive Behaviour Therapy for the Prevention, Treatment and Relapse Prevention of Eating Disorders: A Systematic Review

**DOI:** 10.1101/2022.07.15.22277685

**Authors:** Nilima Hamid

## Abstract

**Background and Objectives:** Eating disorders (EDs) are undertreated worldwide. In the UK the lag between recognition of symptoms and treatment ranges from about 15 months to in excess of two years. Internet-based Cognitive Behaviour Therapy (ICBT) could be a viable alternative to face-to-face Cognitive Behaviour Therapy (CBT) that avoids the negative impacts of delayed interventions. Based on evidence from randomised controlled trials (RCTs) this systematic review investigated the effectiveness of minimally guided self-help ICBT, without face-to-face therapy, for the prevention, treatment and relapse prevention of all types of EDs in adults.

**Methods:** The electronic databases MEDLINE, PsychINFO, CENTRAL, Scopus, and Web of Science were searched between 1991 to 2021. Inclusion criteria specified RCTs with ICBT versus inactive comparison groups. The Cochrane Risk of Bias Tool was used for quality assessments. Qualitative synthesis and meta-analyses were conducted.

**Results:** Findings showed medium and large significant beneficial effect sizes for the prevention and treatment studies, respectively, whereas relapse prevention yielded mainly small non-significant beneficial effect sizes. Only the treatment studies reached clinical significance and cognitive symptoms improved more than behavioural symptoms.

**Conclusions:** This systematic review reinforces the vital need to provide evidence-based Internet interventions at times when face-to-face treatment is not an option as has been the case during the COVID-19 pandemic. Although ICBT is a promising intervention for eating disorders in adults and may be more effective than face-to-face CBT for treating cognitive symptoms further high-quality ED RCTs are required to increase the evidence-base and enable more precise meta-analyses to reach definitive conclusions.

**Highlights:**

- Findings showed medium and large significant beneficial effect sizes for the prevention and treatment studies, respectively, whereas relapse prevention yielded mainly small non-significant beneficial effect sizes.
- For the treatment studies, 35% of effect estimates were clinically important. None of the effect estimates for either the prevention or the relapse prevention studies reached clinical significance.
- There were statistically significant improvements on comorbid depression and anxiety for the treatment programmes.
- Cognitive symptoms improved more than behavioural symptoms and it is suggested that ICBT may be more effective than face-to-face CBT for treating cognitive symptoms.

## INTRODUCTION

Eating disorders (EDs) are undertreated worldwide (1). In the UK the lag between recognition of symptoms and treatment ranges from about 15 months to in excess of two years (2). Face-to-face Cognitive Behaviour Therapy (CBT) is the primary treatment for EDs in adults but its availability is limited and Internet-based Cognitive Behaviour Therapy (ICBT) could provide a cost-effective evidence-based alternative to face-to-face CBT that avoids the negative impacts of delayed interventions.

It is proposed that ICBT ED programmes could reduce the financial burden to the healthcare system, patients, their families, and facilitate early intervention mitigating the disruptions to social functioning, education and employment prospects of individuals with an ED, improving their overall quality of life. At the moment, due to being a relatively recent approach high-quality research on ICBT for EDs is minimal. It is important to ascertain both short and long-term effectiveness, cost-effectiveness, patient safety and acceptability of ICBT to support the dissemination and implementation of these programmes and to develop them further to maximise benefits to patients. Currently, the COVID-19 pandemic has increased the risk of developing an ED and exacerbated existing EDs in at risk groups globally (3). Thus, leading to a demand for remote interventions such as ICBT during a period of limited access to face-to-face healthcare services.

Based on evidence from randomised controlled trials (RCTs) this systematic review investigated the effectiveness of minimally guided self-help ICBT, without face-to-face therapy, for the prevention, treatment and relapse prevention of all types of EDs in adults.

## METHODS

This systematic review was conducted in accordance with the Preferred Reporting Items for Systematic Reviews and Meta-Analyses (PRISMA) guidelines (4). The electronic databases MEDLINE, PsychINFO, CENTRAL, Scopus, and Web of Science were searched between 1991-2017 with an updated search to 2021. Inclusion criteria specified RCTs with ICBT versus inactive comparison groups (i.e., waiting-list/delayed-treatment or treatment-as-usual controls) and adult participants. Inactive comparisons were used because of the lack of sufficient studies investigating ICBT versus alternative active comparisons. Only RCTs published in peer-reviewed journals were considered - not study protocols, nor grey literature (e.g., unpublished manuscripts, conference abstracts or government publications). There were no geographical or gender restrictions, but non-English language reports were excluded.

The Cochrane Risk of Bias Tool was used for quality assessments (5). Following qualitative synthesis, random effects meta-analyses were conducted using Review Manager 5.3 (6). Standardised Mean Difference (SMD) in this case, Hedges’ (adjusted) *g* (5) was the (post-treatment between group) effect size used in the meta-analyses and are interpreted according to the thresholds of 0.2 = small effect size, 0.5 = medium effect size, and 0.8 = large effect size (7). I^2^ which is the extent of heterogeneity (inconsistency between studies, 0% to 100%) was calculated, and the strength of the whole body of evidence was assessed using the Grades of Recommendation, Assessment, Development and Evaluation (GRADE) approach (5).

The keywords searched in the titles and abstracts for each database are shown in Table 1.

**TABLE 1.**
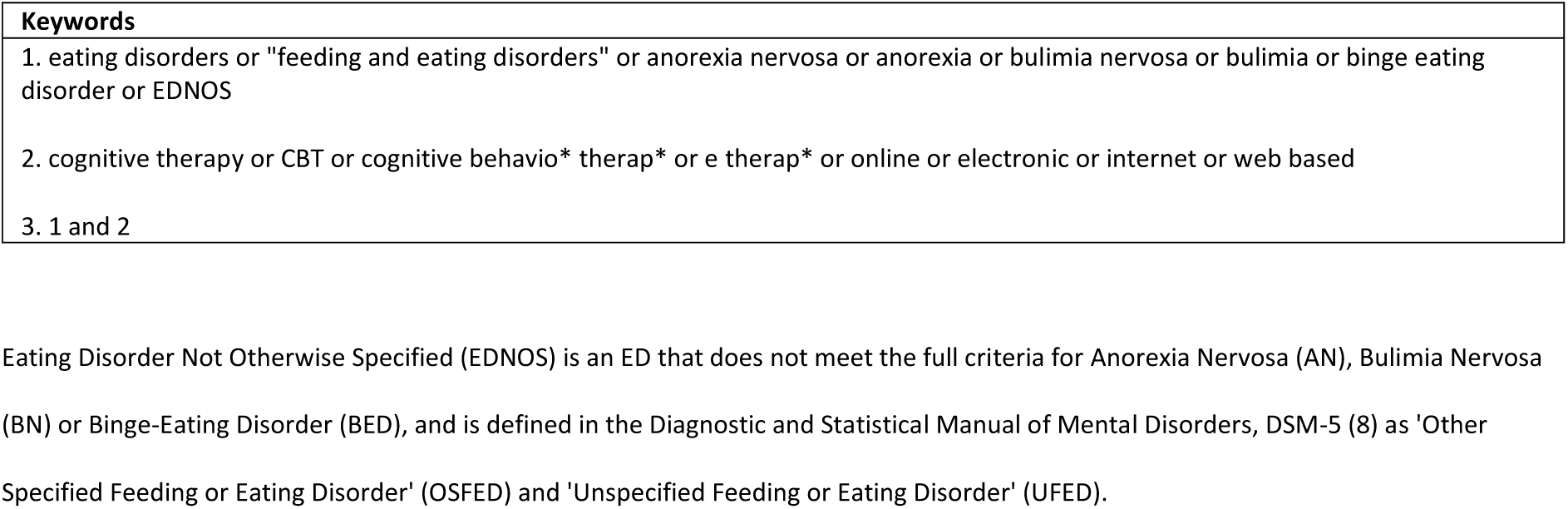
Search Strategy for Databases.

## RESULTS

Fourteen RCT studies presented in 16 reports were included in this systematic review (*N* = 2076). Figure 1 describes the study selection process.

**FIGURE 1.**
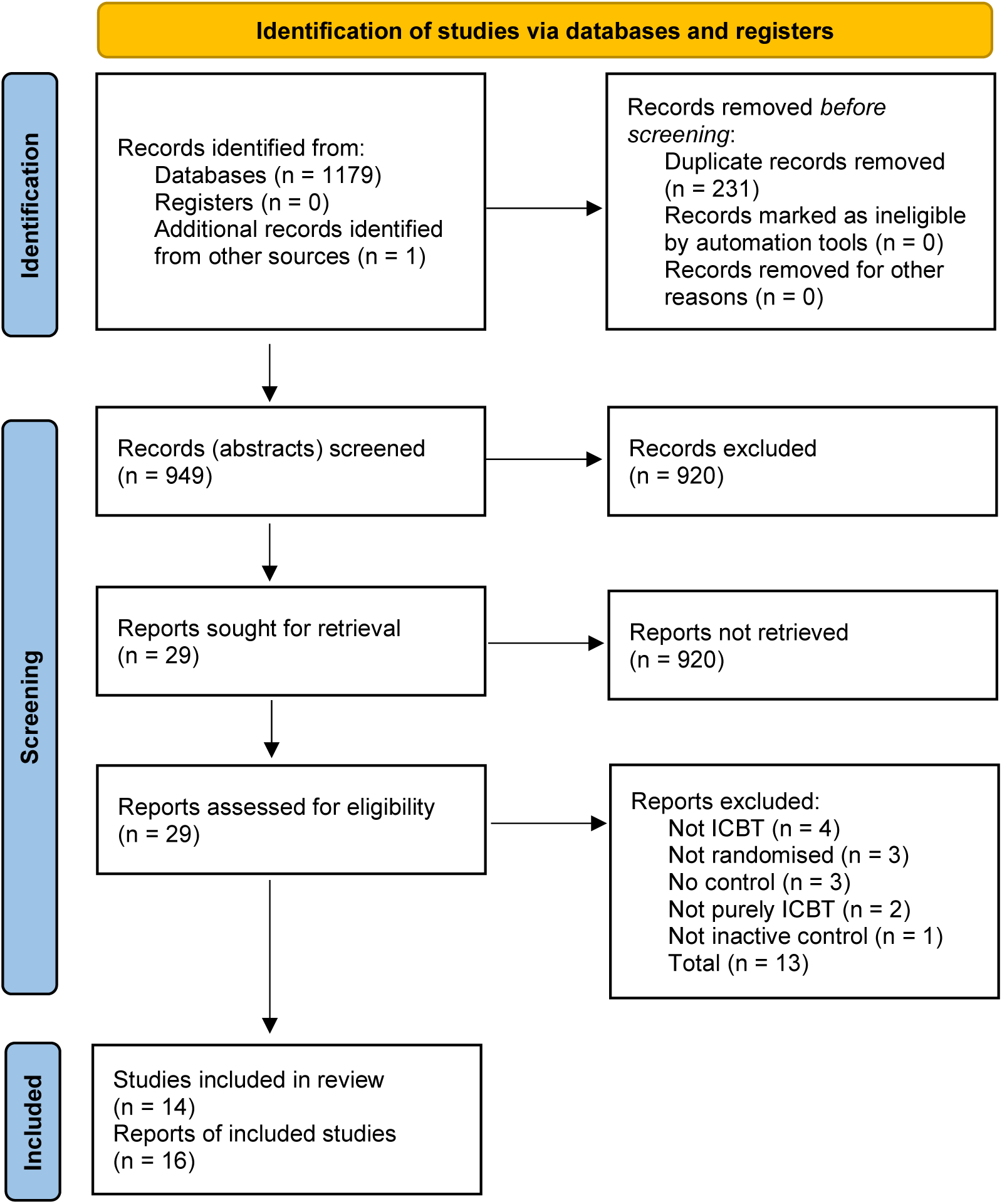
PRISMA Flow Diagram (9)

Quality assessments using the Cochrane Risk of Bias Tool (5) identified three out of 14 RCTs to be of very low methodological quality, whilst six were of low quality and five of 14 were rated as moderate to high quality.

Included studies are listed in Table 2. The ICBT ED prevention, treatment and relapse prevention programmes are interactive, multimedia, online interventions based on CBT principles with modules consisting of psychoeducation, cognitive restructuring, behavioural modification and relapse prevention, all reinforced by homework assignments and email support. Prevention studies also included an asynchronous discussion group and relapse prevention studies offered real-time online chat sessions.

**TABLE 2.** Study Characteristics.

| Study | Intervention | Control |
| --- | --- | --- |
| **Prevention Studies** |  |  |
| Jacobi et al. (10) | *Student Bodies* (SB) adapted for a German population | WLC |
| Jacobi et al. (11); Volker et al. (12) | *Student Bodies* adapted for women with subthreshold ED (SB+) | WLC |
| Low et al. (13) | *Student Bodies* with/without moderated discussion group/no discussion group | WLC |
| Saekow et al. (14) | *Student Bodies* adapted to target restrictive eating, bulimic and compensatory behaviour (SB-ED) | DTC |
| Taylor et al. (15) | *Student Bodies* | DTC |
| Taylor et al. (16) | Image and Mood (IaM) adapted from *Student Bodies* | DTC |
| Winzelberg et al. (17) | *Student Bodies* | DTC |
| Zabinski et al. (18) | *Student Bodies* | DTC |
| **Treatment Studies** |  |  |
| Carrard et al. (19) | ICBT programme SALUT-BED adapted from the SALUT-BN programme | DTC |
| Ruwaard et al. (20) | ICBT programme for BN (Interapy) or Bibliotherapy (unguided) | DTC |
| Sánchez-Ortiz et al. (21) | ICBT programme Overcoming Bulimia Online (OBO) | DTC |
| Wagner et al. (22) | ICBT programme for BED (ICBT-BED) | DTC |
| **Relapse Prevention Studies** |  |  |
| Fichter et al. (23); Fichter et al. (24) | ICBT programme (VIA) for relapse prevention of AN | TAU |
| Jacobi et al. (25) | ICBT programme (IN@) for relapse prevention of BN | TAU |

WLC: waiting-list control, DTC: delayed-treatment control, TAU: treatment-as-usual.

Studies were conducted in the UK, USA and mainland Europe. Most participants were female. The prevention studies included young college or university students recruited via Higher Education institutions, but in the treatment and relapse prevention studies the participants were slightly older clinical samples, except for one treatment study which used a student population. The duration of prevention studies were 8-10 weeks, whereas treatment and relapse prevention studies were longer (3-9 months), highlighting that early intervention would benefit both patients and the healthcare system.

The studies were consistent in their use of outcome measures well known in the EDs field, which are standardised, reliable and valid instruments.

### Meta-Analyses

Statistically significant effect sizes (p ≤ 0.05), extent of heterogeneity (I^2^), and GRADE ratings for the included studies are presented in Table 3.

**TABLE 3.** Meta-Analyses: statistically significant effect sizes.

| Prevention Studies: At-Risk Patients |  |  |
| --- | --- | --- |
| Outcome | Effect Size<br>SMD = *g* (95% CI) I<sup>2</sup> % | GRADE Rating |
| EDE-Q |  |  |
| Eating Disorder |  |  |
| Cognitive Symptoms |  |  |
| Restraint | -0.41 (-0.80, -0.02) 75 | Very Low |
| Weight concern | -0.31 (-0.57, -0.06) 50 | Very Low |
| Global score | -0.46 (-0.82, -0.09) 79 | Very Low |
| EDI/EDI (2) |  |  |
| Drive for thinness | -0.45 (-0.61, -0.28) 26 | Low |
| WCS | -0.47 (-0.82, -0.11) 80 | Very Low |
| Treatment Studies: [Bulimia Nervosa (BN) and Binge-Eating Disorder (BED) Patients] |  |  |
| Outcome | Effect Size<br>SMD = *g* (95% CI) I<sup>2</sup> % | GRADE Rating |
| EDE-Q |  |  |
| Eating Disorder |  |  |
| Cognitive Symptoms |  |  |
| Eating concern | -0.99 (-1.34, -0.63) NA | Very Low |
| Weight concern | -1.11 (-1.47, -0.75) NA | Very Low |
| Shape concern | -0.65 (-1.29, -0.01) 80 | Very Low |
| Global score | -0.70 (-1.22, -0.17) 77 | Very Low |
| EDE-Q |  |  |
| Behavioural Items |  |  |
| Binge rate | -0.66 (-1.11, -0.22) 69 | Very Low |
| EDI/EDI (2) |  |  |
| Bulimia | -0.85 (-1.33, -0.37) NA | Very Low |
| Interoceptive awareness | -0.51 (-0.97, -0.05) NA | Very Low |

| BDI/CES-D | **-0.47 (-0.74, -0.20)** 0 | Very Low |
| --- | --- | --- |
| SCL-90R | **-0.30 (-0.57, -0.03)** 0 | Very Low |
| **Relapse Prevention Study: [Anorexia Nervosa (AN) and related Eating Disorder Not Otherwise Specified (EDNOS) Patients]** |  |  |
| **Outcome** | **Effect Size**<br>SMD = *g* (95% CI) *I*<sup>2</sup> % | **GRADE Rating** |
| EDI/EDI (2) |  |  |
| Maturity fears | **-0.43 (-0.70, -0.16)** NA | Low |
| Social insecurity | **-0.29 (-0.56, -0.03)** NA | Low |

SMD = Standardised Mean Difference, in this case, Hedges' (adjusted) *g*; 95% CI = 95% confidence interval; *I*<sup>2</sup> = extent of heterogeneity (inconsistency between studies); EDE-Q (Eating Disorders Examination Questionnaire) (26); EDI/EDI-2 (Eating Disorders Inventory) (27); WCS (Weight Concern Scale) (28); BDI/BDI-2 (Beck Depression Inventory) (29); CES-D (Centre for Epidemiological Studies Depression Scale) (30); SCL-90R (GSI) (Symptom Checklist 90R, General Symptom Index Score – general psychopathology/anxiety) (31); Grades of Recommendation, Assessment, Development and Evaluation (GRADE) Ratings; NA = Not Applicable.

These results are limited by the small number of included studies. There was a pattern of very high heterogeneity (I^2^) across the meta-analyses, thus lowering confidence in the combined results. The GRADE ratings for the outcomes were mostly very low and decrease the credibility of the results.Analyses undertaken reinforced that publication bias was not of great concern in this study and the search strategy was exhaustive, but searching grey literature would have increased confidence in the results.

## DISCUSSION

### Is ICBT Effective?

This systematic review evaluated the effectiveness of ICBT interventions for all types of EDs in adults and found medium and large significant beneficial effect sizes for the prevention and treatment studies, respectively, whereas relapse prevention yielded mainly small non-significant beneficial effect sizes.

In the treatment studies, 35% of effect estimates exceeded the minimally important difference (MID) SMD = 0.5 (32); thus, these results were clinically significant and were evident for *the Eating Disorders Examination-Questionnaire’s* (EDE-Q) cognitive symptoms (eating, weight, and shape concerns, and global score); the EDE-Q’s behavioural item (binge rate); *the Eating Disorders Inventory’s* (EDI) bulimia (binge-eating and purging behaviour) and interoceptive awareness (sensations of hunger and satiety). None of the effect estimates for either the prevention or the relapse prevention studies reached clinical significance. This could be due to the greater scope for improvement in the participants of the treatment studies.

The moderate favourable influence on depression and a small positive impact on general psychopathology/anxiety for the treatment programmes are important as EDs are frequently comorbid with depression or anxiety (2).

Internet-based CBT influenced cognitive symptoms (e.g., eating and weight concerns) more than it affected behavioural symptoms (e.g., binge or purge rates). By contrasting the effect sizes of this review with those provided by Linardon et al. (33) it is suggested that ICBT may be more effective than face-to-face CBT for treating cognitive symptoms. This may be because participants have more time to absorb the cognitive content in ICBT than in face-to-face CBT. Improvements in both behavioural and cognitive symptoms are imperative as residual cognitive symptoms may lead to relapse (33). If the impact on cognitive symptoms leads to lower relapse rates, then the long-term effectiveness of ICBT would be significantly more effective than other treatments. While direct comparisons between ICBT and face-to-face CBT are needed, these findings are very encouraging.

For the ICBT ED prevention programmes, the evidence-base that supports ICBT’s effectiveness, although it varies in quality and is moderate in size, still provides reasonable confidence in these programmes. This is in spite of the use of programme completers’ data in the meta-analyses, which may present biased over-estimates. The very large effect sizes found for the treatment programmes is countered by the varied quality and limited evidence-base. Even though one of the two relapse prevention programmes is of reasonably high quality, the evidence-base for these programmes is very small. It is difficult to reach conclusions about the treatment and relapse prevention programmes in spite of the fact that the more cautious intention-to-treat (ITT) results were used in the meta-analyses. Many more high quality ICBT ED RCTs are needed to enable precise meta-analyses and robust conclusions.

Internet-based CBT for prevention is a difficult issue for dissemination as most individuals remain at some risk after prevention trials and resources are simply not available for people to then go to a higher level of care, like face-to-face. Nevertheless, this review provides preliminary evidence that ICBT interventions could be offered to adult community and student populations in Western countries in an adaptive stepped care approach and is likely to be cost-effective compared to face-to-face CBT. Such an adaptive stepped care approach would be more flexible and tailored to individual needs. Patients could then be monitored closely and receive more intensive support earlier or when indicated.

A number of meta-analyses have investigated studies under the prevention category (e.g., Harrer et al. (34)) and a few have looked at treatment and relapse prevention studies (e.g., Aardoom et al. (35); Schlegl et al. (36)) but these studies did not focus entirely on ICBT. Further, Taylor et al. (37) provides a broad summary of the evidence supporting the effectiveness of Internet interventions and describe lower effect sizes for EDs and substance abuse compared to depression and anxiety.After this review was completed a cluster randomised RCT of ICBT versus usual care for women with binge-purge EDs was found which showed significant reduction in eating disorder psychopathology in the intervention group compared to control at postintervention and at follow-up (38). Another study is more problematic as it has not been published but does address a core issue of the review: Jacobi et al. (39) – RCT analysing indicated, Internet-based prevention for women with anorexia nervosa (AN) symptoms.

At the time of initiation, this review updated the Loucas et al. (40) study and included five new studies. Subsequently, Linardon et al. (41) published their study but did not include relapse prevention studies and looked at e-therapies more broadly rather than focussing on ICBT. In agreement with this systematic review, these two latest and most comprehensive systematic reviews on the effectiveness of e-mental health ED programmes have taken an optimistic perspective, while clarifying the need for further research. Interestingly, larger effect sizes were found in the current review compared to these two reviews. This could be due to focusing exclusively on RCTs and Internet-delivered CBT and indicating that at this time ICBT may be more effective for treating EDs than other forms of Internet psychotherapies and delivery via downloadable software, CD-ROMs, and mobile applications. The strengths of this review are arguably the inclusion of the best-evidence base (just RCTs), specifically ICBT programmes, all types of EDs and solely adult participants. Notably, these characteristics are not addressed together in previous reviews and as such this review likely provides clearer estimates of the effectiveness of ICBT for EDs in adults.

### Future Research

More RCTs, particularly treatment and relapse prevention trials, are needed which accord with the CONSORT statement (42) and aligned with the research by Proudfoot and colleagues (43) researchers should follow the guidance for conducting and reporting ICBT research. Such research will facilitate more precise meta-analyses to reach unequivocal conclusions. Replication of the large effect sizes demonstrated for the treatment programmes would support the necessity of developing these programmes further. Including non-English language studies could have retrieved RCTs from populations that do not primarily speak English (e.g., individuals living in Asia, Africa, South America). However, at this time the generalisability of ICBT programmes included in this systematic review is limited predominantly to young Caucasian women in Western countries. Further research is essential to confirm whether the findings can be transferred to other ethnicities, countries beyond the UK, USA and mainland Europe, and different healthcare systems.

Finally, the use of digital technology (computers, tablets, mobile phones) is prevalent worldwide and has the potential to extend the reach of mental health interventions even in low- and middle-income countries (44). As Taylor et al. (45) outline, the COVID-19 pandemic has led to a shift in using telehealth instead of face-to-face consultations and highlighted the need for digital mental health interventions to become part of routine care. Barriers to digital services (e.g., training of therapists, licensing regulations, patient safety concerns, ensuring the privacy of patients, reimbursement for digital therapies, and further research into the gaps in knowledge about digital therapies) need to be addressed, and it is necessary to exploit new technologies, including virtual/augmented reality, to optimise the content and delivery of ICBT ED programmes with the goal of providing timely, flexible, personalised, interactive, cost-effective, and evidence-based interventions.

## Conclusions

In the midst of the COVID-19 pandemic, results from this systematic review reinforce the vital need to provide evidence-based Internet interventions at times when face-to-face treatment is not an option. Minimally guided self-help ICBT programmes, without face-to-face therapy, have shown improvements in ED risk factors, onset, symptoms and relapse in adults. Internet-based CBT is promising for the prevention, treatment and relapse prevention of EDs, but further high-quality RCT research is necessary to reach more definite conclusions.

## Supporting information

Research Reporting Checklist

## Data Availability

The data that support the findings of this study are available from the corresponding author upon reasonable request.

## Ethical Considerations

This work was secondary research (a systematic review) and did not involve the use of human subjects thus there were no ethical considerations.

## Acknowledgements

This journal article is based on the dissertation submission for the award of MSc in Psychiatry (Cardiff University) by Nilima Hamid.

